# Risdiplam in adult non-sitter patients with 5q spinal muscular atrophy: a non-interventional, single centre, observational cohort study

**DOI:** 10.1101/2022.06.20.22276486

**Authors:** Nancy Carolina Ñungo Garzón, Inmaculada Pitarch Castellano, Teresa Sevilla, Juan F Vázquez-Costa

## Abstract

**Objective:** To describe the safety and efficacy of risdiplam in non-sitter adult patients with 5q spinal muscular atrophy (SMA).

**Methods:** Type 2 SMA adult patients, who were not eligible for nusinersen, were offered risdiplam through the expanded access program. Patients were followed up with a battery of scales and clinical measures.

**Results:** Six non-sitter patients (17 – 46 years old) were treated with risdiplam. One patient reported mild adverse events (dyspepsia and headache). After one year of treatment, all patients showed clinically meaningful improvements in at least one scale and none of them showed any clinically meaningful deterioration. Two patients showed a clinically significant increase in the body mass index and other two in the revised upper limb module. Moreover, clinically meaningful improvements were found in motor (axial and upper limbs), bulbar (speech and swallowing) and respiratory (coughing) domains of functional scales, in five patients. Four subjects achieved at least one of the goals set with the goal attainment scale (GAS).

**Discussion:** This series suggests the safety and efficacy of risdiplam in non-sitter adult SMA patients. In these patients, functional scales and GAS are more appropriate than motor scales to detect changes, because they include axial, bulbar and respiratory domains.

## Introduction

Risdiplam is an oral medication approved for the treatment of patients with spinal muscular atrophy (SMA) type 1, 2 and 3 in the USA and Europe[1]. It modifies the SMN2 (survivor neuronal motor 2) pre-mRNA splicing to promote the inclusion of exon 7 and increase the production of functional SMN protein[2]. Its approval for type 2 and 3 patients was based on the results of the SUNFISH study part 2, which showed efficacy in non-ambulant patients, including a small subgroup of adults[3]. However, non-sitters were excluded from this study [3]. Due to their complex spines, non-sitter patients cannot be treated with nusinersen [4], an intrathecally administered antisense oligonucleotide also approved for the treatment of SMA patients. Consequently, the efficacy of disease-modifying therapies in non-sitter patients is not well known and they may have limited access to them, despite their bad prognosis [5].

The aim of this study was to assess the safety and efficacy of risdiplam in a series of non-sitter adult SMA patients.

## Methods

Six adult (>16 years old) type 2 patients consecutively visited in Hospital la Fe between October and December 2020 and fulfilling the inclusion and exclusion criteria of the expanded access program (EAP, NCT04256265), were offered risdiplam in a single daily dose of 0.25 mg/kg for < 20 kg and 5 mg for those > 20 kg [3]. For this study, only non-sitter patients (not able to maintain the seated position for 5 seconds, with support of one or both upper limbs) have been considered. In the case patients of patients previously treated with nusinersen, risdiplam was started at least four months after last nusinersen dose. The following outcome measures were prospectively recorded at baseline and after 6 and 12 months of treatment: body mass index (BMI), the percent-predicted forced vital capacity (FVC%), the Revised Upper Limb Module (RULM), the Egen Klassification (EK2), the Revised Amyotrophic Lateral Sclerosis Functional Rating Scale (ALSFRS-R). Moreover, the ALSFRS-R and EK2 scores 12 months before treatment were retrospectively recorded. Clinically meaningful changes for each scale were defined as previously suggested[6] (≥2 points for RULM, EK2 and ALSFRS-R) and, based on our experience and previous literature, a change >5% in the BMI and FVC was also considered meaningful. Three personalized functional goals were established at the baseline visit (Table 1), after discussion with each patient, and re-evaluated after 12 months with the Global Attainment scale (GAS)[7]. The use of assistive devices such as non-invasive ventilation (NIV), cough assist or percutaneous endoscopic gastrostomy (PEG), the patient’s global impression of change (P-GIC) and the clinical impression of global change (C-GIC) were also recorded at the 12-months visit. Safety was evaluated through the monitoring and recording of adverse events (AE) and laboratory assessments at 1, 6 and 12 moths.

**Table 1.** Demographical and baseline clinical characteristics of SMA patients (A) and quantification of change in scores (Δ) between the baseline and the 12-months visits (B). In bold, clinically meaningful results. ALSFRS-R: Revised version of the Amyotrophic Lateral Sclerosis Functional Scale; BMI: body mass index; C-GIC: clinical global impression of change; EI: Entry Item; EK2: Egen Klassifikation 2; F: Female; FVC%: Percent-predicted Forced Vital Capacity; GAS: Goal Attainment scale; M: Male; P-GIC: patient global impression of change; RULM: Revised Upper Limb Module. EK2: Egen Klassifikation 2; GAS: Goal Attainment Scale; NIV: noninvasive ventilation; RULM: Revised Upper Limb Module; SMA: spinal muscular atrophy; SMN2: survival motor neuron 2 gene.

| Patient N° | 1 | 2 | 3 | 4 | 5 | 6 |
| --- | --- | --- | --- | --- | --- | --- |
| **A. Demographic and clinical characteristics** |  |  |  |  |  |  |
| **Sex** | F | M | M | F | M | F |
| **Age (years)** | 31-40 | 15-20 | 15-20 | 21-30 | 21-30 | 31-40 |
| **SMN2 copy number** | 3 | 3 | 3 | 1 | 3 | 4 |
| **FVC (%)** | NA | NA | NA | 16 | 18 | 34.4 |
| **Ventilatory support** | NIV | NIV | NIV | NIV | No | No |
| **BMI** | 8 | 6.6 | 16.6 | 17.9 | 17.4 | 26.3 |
| **Nutritional Support** | Oral | Oral | No | Oral | No | No |
| **Previous SMA treatment** | No | Nusinersen (9 doses) | No | Nusinersen (3 doses) | No | No |
| **RULM EI** | 0 | 3 | 3 | 1 | 1 | 1 |
| **RULM score** | 0 | 10 | 8 | 0 | 1 | 0 |
| **EK2** | 45 | 28 | 22 | 37 | 30 | 29 |
| **ALSFRS – R** | 11 | 17 | 23 | 13 | 24 | 21 |
| **Goals (GAS)** | 1. Improve swallowing<br>2. Talk without shortness of breath<br>3. Use cutlery alone | 1. Improve head control<br>2. Eating and drinking in the sitting position<br>3. Less shortness of breath | 1. Eating alone<br>2. Improve typing<br>3. Improve head control | 1. Drink independently (with a straw)<br>2. Reduce fatigue<br>3. Improve typing | 1. Eating alone<br>2. Improve speech<br>3. Improve nutrition | 1. Avoid NIV initiation<br>2. Reduce fatigue<br>3. Improve “pincer grip” |
| **B. Change in scores ($\Delta$) after 12 months of treatment** |  |  |  |  |  |  |
| **$\Delta$ BMI (%)** | -2.5 | **12.1** | 0 | -0.6 | **9,8** | -1.1 |
| **$\Delta$ FVC%** | NA | NA | NA | 2 | -2 | 1,5 |
| **$\Delta$ RULM score** | 0 | 1.5 | **3.5** | **3** | 0 | 0 |
| **$\Delta$ EK2** | **6** | **3** | 1 | **7** | **4** | **5** |
| **$\Delta$ ALSFRS – R** | **7** | **2** | 1 | **9** | 1 | 1 |
| **C – GIC** | 1 | 1 | 1 | 1 | 1 | 1 |
| **P - GIC** | 1 | 1 | 0 | 1 | 2 | 1 |
| **GAS** | 0 | **5.9** | 0 | **22.8** | **18.3** | **20.4** |

All patients signed informed consent before the start of risdiplam, and for the publication of this report. Data collection and processing were approved by the Ethics Committee for Biomedical Research of the La Fe Hospital (Valencia).

## Results

Six non-sitter adult SMA patients started treatment with risdiplam. Their baseline demographic and clinical characteristics are summarized in Table 1A and supplementary table. Briefly, 5 patients showed a low BMI (<18.5), 4 patients routinely used NIV, 4 patients showed only minimal residual mobility in upper limbs (RULM entry item ≤ 1) and two patients had been previously treated with nusinersen (one was discontinued due to the lack of lumbar access and the other due to the lack of efficacy).

Regarding the respiratory function, there were no changes in the use of ventilatory support, nor clinically meaningful changes in the FVC%, albeit a valid FVC could not be obtained in half of patients (Table 1B). Despite this, 3 patients reported an improvement in their cough ability in EK2, resulting in a reduction of the cough assist need. At the nutritional level, three patients reported improved swallowing in functional scales, 2 of them showing clinically meaningful improvements in the BMI (Table 1B). Of note, in one of these patients gastrostomy had been proposed due to sustained weight loss in previous visits, but was ruled out after the improvement with risdiplam.

Three patients scored 0 at the baseline RULM. Nonetheless, 2 patients showed clinically meaningful improvements, while the rest remained stable or improved mildly (Table 1B). Conversely, no floor effect was found in EK2 nor in ALSFRS-R. Moreover, five patients showed clinically meaningful improvements in any of their motor (head control and distal upper limbs), bulbar (speech and swallowing) or respiratory (coughing) domains (Table 1A and Figure 1).

**Figure 1.**
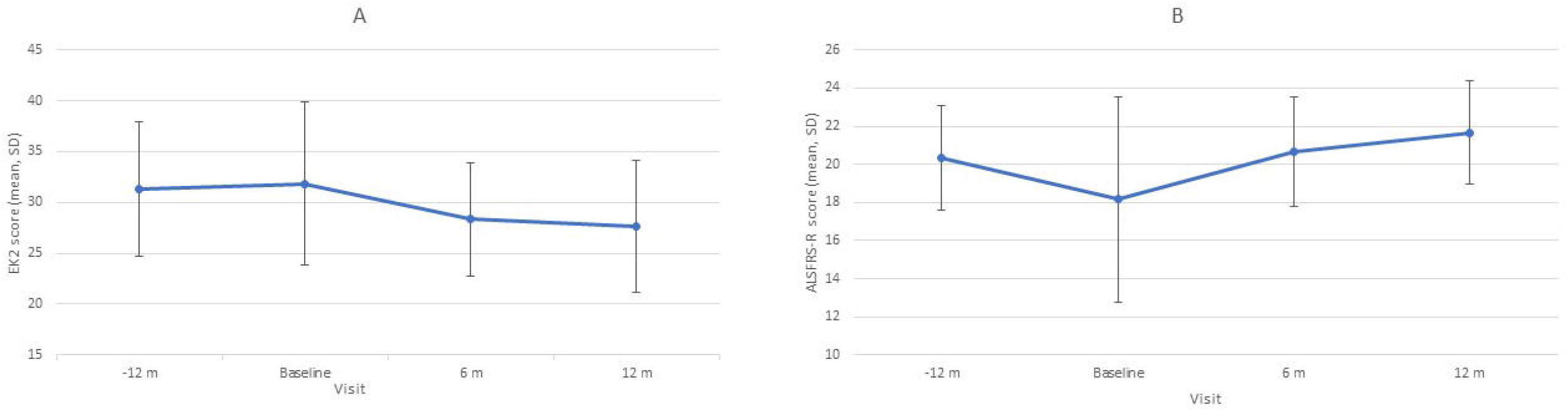
Graphical representation of evolution of total scores on the EK2 (A) and ALSFRS-R (B) scales from 12 months prior to treatment to 12 months after treatment. Of note, better function is represented as a lower score in EK2 and higher in ALSFRS-R. ALSFRS-R: Revised version of the Amyotrophic Lateral Sclerosis Functional Scale; EK2: Egen Klassifikation 2.

Overall, all patients showed clinically meaningful improvements in at least one outcome measure, while none of them showed clinically meaningful deterioration in any of them. Accordingly, the C-GIC scored mild improvements in all patients. Regarding the P-GIC, 4 patients reported mild improvements, 1 a moderate improvement and another did not report change in their global condition. Finally, in the GAS, 2 patients reporting some improvement, did not achieve any of their goals set in the baseline visit, while the other 4 achieved at least one of them (Table 1B).

One patient reported mild AEs (gastrointestinal symptoms and headache) and discontinued the medication after 10.5 months of treatment. However, she experienced a functional deterioration after withdrawal and restarted it at 12 months, without any new AE. No clinically relevant laboratory changes were found in any of the patients.

## Discussion

Here, we describe our experience with risdiplam in non-sitter adult SMA patients, a subgroup of patients for whom safety and efficacy data are lacking.

Overall, risdiplam was well tolerated and there were clinically meaningful improvements in at least one domain in all patients, also suggesting its efficacy. Improvements included motor (head control, distal upper limbs, fatigability), bulbar (speech and swallowing) or respiratory (coughing) domains, which are relevant for non-sitter patients. It is important to note that these domains are either difficult to measure or not measurable at all with most motor scales, which are currently used as the gold standard outcome measures in both clinical trials and clinical practice [5,8].

Specifically, in our series, half of patients could not perform or showed floor effects with both RULM and FVC, which are probably the most widely used measures in non-sitters [5,8]. Consequently, non-sitter adults have been systematically excluded from clinical trials and there is also a lack of real-world data on these patients, resulting in limited access to disease modifying treatments. This study, in line with previous works [8], shows that functional scales (specifically EK2) and the GAS are helpful to set personalized goals and measure individual responses to treatment in adult non-sitter patients, because they can detect subtle but meaningful changes in relevant aspects of the daily life. Hence, regulatory agencies increasingly recommend its use in both clinical trials and practice. The goals set here before treatment show that finger movements, head control, swallowing, breathing and fatigue are the most important issues for non-sitter patients. Remarkably, despite all patients reporting some improvement, no patient reached all goals set at baseline. This could reflect over-ambitious expectations[9]. Nevertheless, one year is probably insufficient time to reach the full response to treatment, as found with nusinersen[10], and it remains still possible for these patients to reach their preestablished goals.

Despite the limitations of this study, mainly the limited sample size, our results suggest the safety and efficacy of risdiplam, and the usefulness of functional scales and GAS for measuring changes in non-sitter adult SMA patients. Larger studies are warranted to confirm the findings of this small series.

## Data Availability

JFVC and NCNG had full access to the database population used to create the study population. All data supporting our findings are available on reasonable request.

## Data availability

JFVC and NCÑG had full access to the database population used to create the study population. All data supporting our findings are available on reasonable request.

## Acknowledgments

We would like to thank Fernando Mora and M Carmen Baviera, for their participation in patients’ assessment, and SMA patients for their participation.

## Study funding

This study has received funding from CUIDAME (PIC188-18) and from Instituto de Salud Carlos III (JR19/00030 PI JFVC). The Centro de Investigación Biomédica en Red de Enfermedades Raras (CIBERER) is initiative from the ISCIII. TS and JFVC are members of the European Reference Network for Rare

Neuromuscular Diseases (ERN EURO-NMD). Sponsors did not participate in the study design, data acquisition and analysis, data interpretation or in writing the article.

## Disclosures

This study has received funding from CUIDAME (PIC188-18).

Dr. Vázquez-Costa is funded by grants of the Instituto de Salud Carlos III (JR19/00030, PI Vázquez), served on advisory boards for Biogen and Roche and received travel and speaker honoraria from Biogen and Roche.

Dr. Pitarch-Castellano served on advisory boards for Avexis and Biogen and received travel and speaker honoraria from Biogen and Roche; principal investigator for ongoing Biogen clinical trial.

## Contribution

NCÑG participated in clinical data acquisition and interpretation and wrote and edited the manuscript.

IPC and TS critically revised the manuscript

JFVC designed the study, participated in clinical data acquisition and interpretation, and wrote and edited the manuscript.

Supplementary table. Clinical data at baseline visit and at 12 months with calculations of change in each parameter. ALSFRS-R: Revised version of the Amyotrophic Lateral Sclerosis Functional Scale; BMI: body mass index; C-GIC: clinical global impression of change; EI RULM: Entry item Revised Upper Limb Module; EK2: Egen Klassifikation 2; FVC%: Percent-predicted Forced Vital Capacity; GAS: Goal Attaintment scale; P-GIC: patient global impression of change; RULM: Revised Upper Limb Module.

## Notes

### Competing Interest Statement

Dr. Vazquez-Costa is funded by grants of the Instituto de Salud Carlos III (JR19/00030, PI Vazquez), served on advisory boards for Biogen and Roche and received travel and speaker honoraria from Biogen and Roche.
Dr. Pitarch-Castellano served on advisory boards for Avexis and Biogen and received travel and speaker honoraria from Biogen and Roche; principal investigator for ongoing Biogen clinical trial.

### Clinical Trial

NCT04256265

